# Distinct Vaccine Efficacy Rates Among Health Care Workers During a COVID-19 Outbreak in Jordan

**DOI:** 10.1101/2022.01.15.22269356

**Authors:** Iyad Sultan, Abdelghani Tbakhi, Osama Abuatta, Sawsan Mubarak, Osama Alsmadi, Adib Edilbi, Ruba Al-Ani, Manar Makhlouf, Rawan Hajir, Omar Khreisat, Majeda A. Al-Ruzzieh, Hikmat Abdelrazeq, Asem Mansour

## Abstract

**BACKGROUND:** We aimed to assess the efficacy of 3 COVID-19 vaccines in a population of health care workers at a tertiary cancer center in Amman, Jordan.

**METHODS:** We evaluated the records of 2855 employees who were fully vaccinated with 1 of 3 different vaccines and those of 140 employees who were not vaccinated. We measured the number of SARS-CoV-2 infections that occurred at least 14 days after the second vaccine dose.

**RESULTS:** The 100-day cumulative incidence of PCR-confirmed SARS-CoV-2 infections was 19.3% ± 3.3% for unvaccinated employees and 1.7% ± 0.27% for fully vaccinated employees. The 100-day cumulative infection rates were 0.7% ± 0.22% in BNT162b2 vaccine recipients (n = 1714), 3.6% ± 0.77% in BBIBP-CorV recipients (n = 680), and 2.3% ± 0.73% in ChAdOx1 recipients (n = 456). We used Cox regression analyses to compare the risks of SARS-CoV-2 infection among the different vaccine recipient groups and found a significantly higher infection risk in BBIBP-CorV (hazard ratio [HR] = 2.9 ± 0.31) and ChAdOx1 recipients (HR = 3.0 ± 0.41) compared to BNT162b2 recipients (*P* = .00039 and .0074, respectively). Vaccinated employees who had no previously confirmed SARS-CoV-2 infections were at a markedly higher risk for breakthrough infections than those who experienced prior infections (HR = 5.7 ± 0.73, *P* = .0178).

**CONCLUSIONS:** Our study offers a real-world example of differential vaccine efficacy among a high-risk population during a national outbreak. We also show the important synergism between a previous SARS-CoV-2 infection and vaccination.

**Funding:** None

## Introduction

By the time of this writing, more than 200 COVID-19 vaccines are available in different stages of clinical development and approval to combat the ongoing pandemic; 19 of them had been approved already.^1^ These vaccines can be generally categorized into 4 groups: (1) inactivated whole virus, (2) protein subunit, (3) mRNA and (4) viral vector vaccines. Approximately 8 billion doses have been administered globally and more than 2 billion people have been fully vaccinated. Real-world examples support the findings of the initial trials of vaccine efficacy, although variable protection is conferred for emerging strains.^2^

Jordan suffered from both the health and financial consequences of the pandemic. By December 5, 2021, 971401 COVID-19 cases have been confirmed, and 11715 deaths have been reported in Jordan. A national vaccination campaign was launched in January 2021 despite anticipated vaccine hesitancy.^3^ More than 3.7 million people have already received 2 doses, representing approximately one-third of Jordan’s population. Vaccines were offered for free to all residents, regardless of nationality. Vaccine recipients, including health care workers (HCWs), were not allowed to choose vaccine manufacturers because the availability of specific vaccine brands was not guaranteed. Shortly after launching the vaccination campaign, the country suffered from a new wave of COVID-19, which was caused predominantly by the B.1.1.7 (i.e. alpha) SARS-CoV-2 variant, according to our institutional random sampling and sequencing data (data not published).

The King Hussein Cancer Center (KHCC) is a tertiary cancer center with approximately 3200 employees and 340 beds. Our Human Resources department maintains vaccination records of all employees. Those with reported close contact to COVID-19 cases, those with respiratory symptoms and those working in high-risk areas were offered free testing, as needed. Additionally, HCWs who had confirmed positive COVID-19 tests performed outside of KHCC (e.g. surveillance for travel or high-risk contacts) were also traced by our Infection Control Program.

The distribution of vaccines, availability of testing and the coincidence of a national outbreak provided an opportunity to compare the effectiveness of different vaccines among KHCC HCWs. In particular, we were able to compare the efficacy of inactivated whole-virus vaccines with that of mRNA and viral vector vaccines. Comparing and reporting the efficacy of these different vaccine types may help policy makers develop best practices and strategies to mitigate future outbreaks.

## Methods

We conducted a cross-sectional study evaluating the vaccination status of our HCWs as of September 30, 2021, along with a retrospective review of previous PCR-confirmed SARS-CoV-2 infections. We considered these data as timed events, with a 0 time-point of 14 days after receiving the second vaccine dose and January 15, 2021, for unvaccinated HCWs. All HCWs employed by KHCC on September 30, 2021, who had been employed for the past 6 months were included in this study. Our endpoint measures comprised the cumulative risk of PCR-confirmed SARS-CoV-2 infections, which is defined by the Jordan Ministry of Health as a positive real-time quantitative RT-PCR cycle threshold of 36. We also assessed the impact of previous SARS-CoV-2 infection and vaccine type on infection incidence with Cox regression analyses and reported the results of this model with hazard ratios (HRs) and *P* values (*P* ≤ .05 considered significant). Vaccine efficacy was calculated using (1-Relative Risk)X100% formula; RR was calculated by dividing the risk of getting an infection among vaccinated group by unvaccinated group over the first 100 days after the start point.^4^ All statistical analyses were performed with R software (v4.0.2). The number of Jordanian residents who were infected with SARS-CoV-2 was obtained by using the COVID-19 package in R.^5^

After the vaccination campaign in Jordan was launched in January 2021, Jordanian citizens were offered free vaccinations and HCWs who refused vaccination were required by law to present valid negative PCR tests twice a week. A vaccination campaign was also launched at KHCC to ease access for busy HCWs and PCR tests were offered for free. Additionally, HCWs were not permitted to choose their administered vaccine type.

After obtaining Institutional Review Board approval (KHCC-IRB#21KHCC110), we accessed deidentified employee records that were maintained by our Human Resources department, which included vaccination records and history of confirmed previous SARS-CoV-2 infections. These data were electronically linked to KHCC laboratory results. The resultant database included the types and dates of vaccine administration, as well as PCR-confirmed SARS-CoV-2 infections.

## Results

By September 30, 2021, we identified 3089 HCWs to be included in our analysis. Their mean age was 34.1 years (standard deviation: 9.5 years). Of the total number of HCWs included in our study, 2855 (92%) were fully vaccinated (i.e. ≥ 14 days post second dose) and 140 (4.5%) had not received any vaccine dose (Table 1). The remaining 94 (3%) HCWs had received either only 1 dose or 2 doses but within the 14-day cutoff period and were therefore not included in our analysis.

**Table 1.** Characteristics of COVID-19 Vaccinations in Health Care Workers at King Hussein Cancer Center in Amman, Jordan.

| Characteristics | No. (% of total) | 100-day events (%) | Hazard ratio | P value |
| --- | --- | --- | --- | --- |
| Total no. | 3089 (100) | 3.2 ± 0.44 | | |
| Fully vaccinated* | 2855 (92) | 1.7 ± 0.27 | – | |
| Not vaccinated | 140 (4.5) | 19.3 ± 3.3 | 8.6 ± 0.23 | <.0001 |
| Fully vaccinated with BNT162b2 | 1714 (55) | 0.7 ± 0.22 | – | |
| Fully vaccinated with BBIBP-CorV | 680 (22) | 3.6 ± 0.77 | 2.9 ± 0.31 | .00039 |
| Fully vaccinated with ChAdOx1 | 456 (15) | 2.3 ± 0.73 | 3.0 ± 0.41 | .0074 |
| Fully vaccinated with Gam-COVID-Vac | 5 (0.2) | 25 ± 22 | NA | NA |
| Previous infection in fully vaccinated | 945 (31) | 0.24 ± 0.17 | – | |
| No previous infection in fully vaccinated | 1910 (62) | 2.4 ± 0.0045 | 5.7 ± 0.73 | .0178 |

\*Fully vaccinated, 14 days post second dose by August 31, 2021.
NA, calculation not available because of small number of participants or short/inadequate follow-up period

Among the fully vaccinated HCWs, 1714 (55% of 3089) received the BNT162b2 vaccine, 680 (22%) received the BBIBP-CorV vaccine, 456 (15%) received the ChAdOx1 vaccine and 5 (0.2%) received the Gam-COVID-Vac vaccine (Table 1). We recorded 274281 follow-up person-days by the study cutoff date, which we distributed into 143939 person-days for BNT162b2, 70645 person-days for BBIBP-CorV, 27263 person-days for ChadOx1 and 514 person-days for Gam-COVID-Vac. The number of HCWs vaccinated per month and the cumulative percentages of HCWs vaccinated are illustrated in figure 1. From March to May 2021, more than 75% of HCWs received their first vaccination, which coincided with our institutional campaign that selectively used BNT162b2. As the number of Gam-COVID-Vac recipients was small, no further analysis was provided for this group.

**Fig 1.**
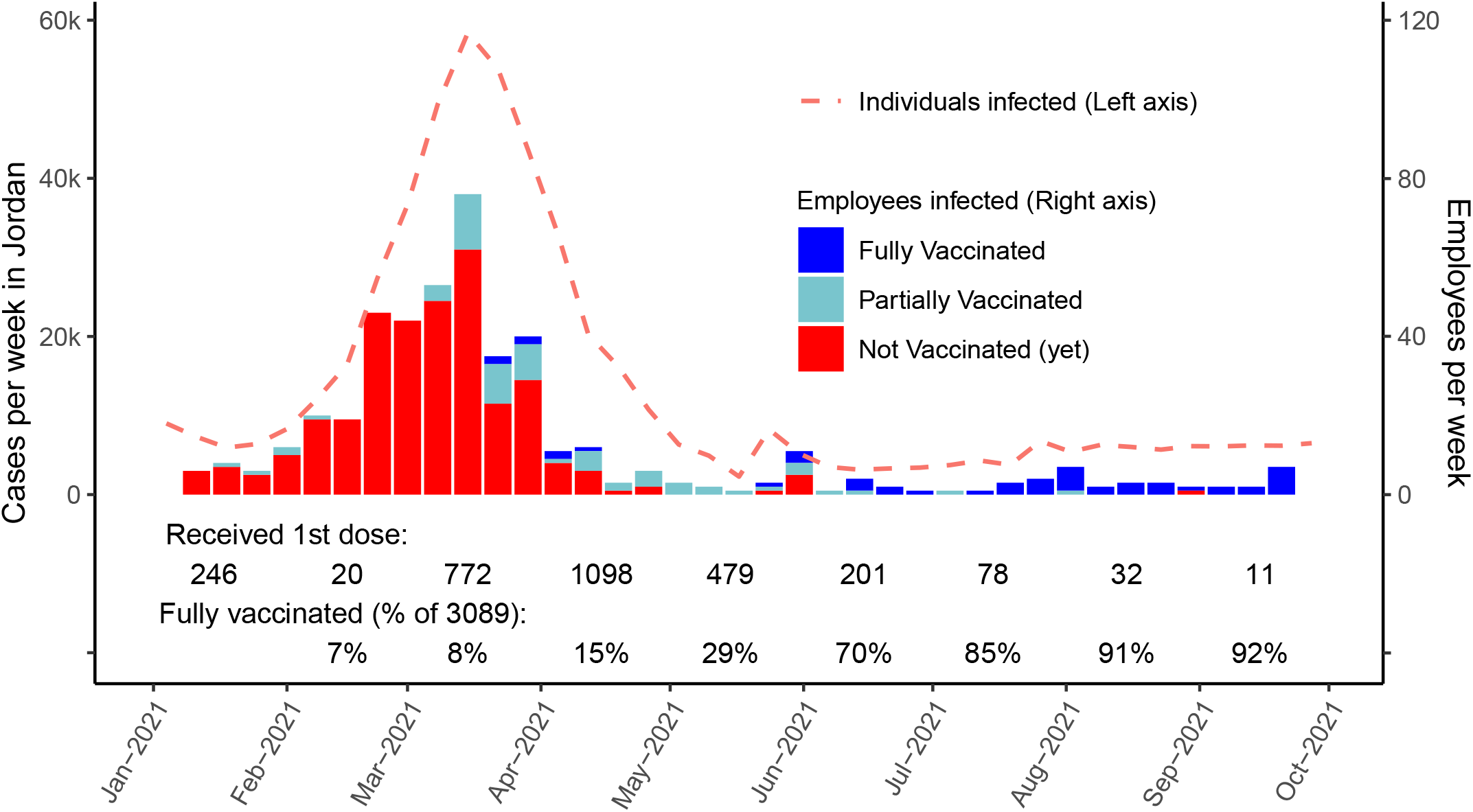
Number of SARS-CoV-2 Infections among Health care workers at King Hussein Cancer Center (bars) with national new cases (dashed line); the last 2 rows show the numbers of individuals who received their first vaccine dose and the cumulative percentage of fully vaccinated employees.

According to our institutional records, 474 documented SARS-CoV-2 infections occurred among our employees during the study period. Of those infections, 353 occurred in HCWs who were not vaccinated, including 28 infections in the 140 employees who were not vaccinated before the study cutoff date and 322 infections in HCWs who were later vaccinated during the study period. An additional 68 infections occurred in partially vaccinated HCWs and 53 infections occurred in HCWs who were fully vaccinated.

The relative risks of getting an infection in the first 100 days post full vaccination for BNT162b2 and BBIBP-CorV relative to unvaccinated employees were 0.04 and 0.23, respectively, yielding an efficacy of 96% and 77%, respectively. The HR of contracting COVID-19 for unvaccinated HCWs versus that of fully vaccinated HCWs was 8.6 (± 0.23) (Table, Figure 2A). Among fully vaccinated HCWs, the risk of contracting COVID-19 was significantly higher for those who received the BBIBP-CorV vaccine, with a cumulative risk of 6.2 ± 1.5 at the study cutoff date (187 days) (Figure 2B). The HR for recipients of BBIBP-CorV versus BNT162b2 was 2.9 ± 0.31 (*P* = .00039) (Table 1). Among vaccinated HCWs with no previous SARS-CoV-2 infection, the HR for breakthrough SARS-CoV-2 infection post vaccination versus those who had experienced previous infection was 5.7 ± 0.73 (*P* = .0178) (Figure 2C). This finding was related to the type of vaccine received, with those vaccinated with BBIBP-CorV and no previous infection having the highest risk.

**Fig 2.**
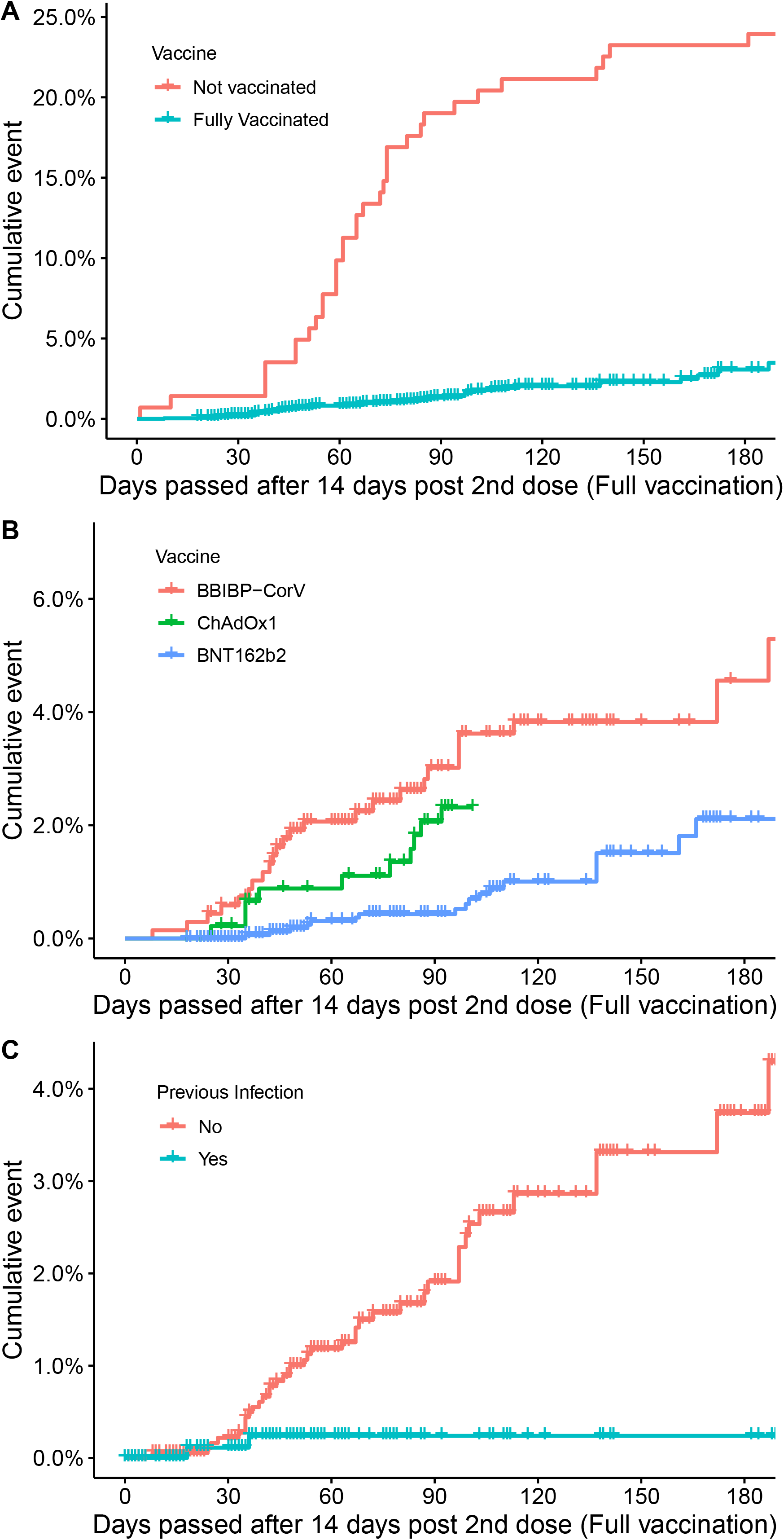
Cumulative SARS-CoV-2 Infections in 2995 Health Care Workers, according to vaccination status (A), vaccine type (B) and having previous infections among vaccinated individuals (C); Gam-COVID-Vac curve not shown on B.

We contacted 52 HCWs who answered questions regarding their symptoms. Thirty-two worked directly with patients and 8 had chronic illnesses, most commonly hypertension (n = 5) and diabetes (n = 3). All individuals reported symptoms after becoming infected. The most commonly reported symptoms were fatigue, headache and loss of taste and/or smell (Figure 3A). Two-thirds (n = 35, 67%) of the HCWs in our study had symptoms that persisted for more than 1 month after infection, which most commonly comprised loss of taste and/or smell, fatigue and concentration difficulty (Figure 3B). Out of all surveyed employees, 4 needed supplemental oxygen, 2 of them had received BBIBP-CorV and 2 had BNT162b2. Only 2 patients were hospitalized, both received had BBIBP-CorV, and none required intensive care admission and/or intubation.

**Figure 3.**
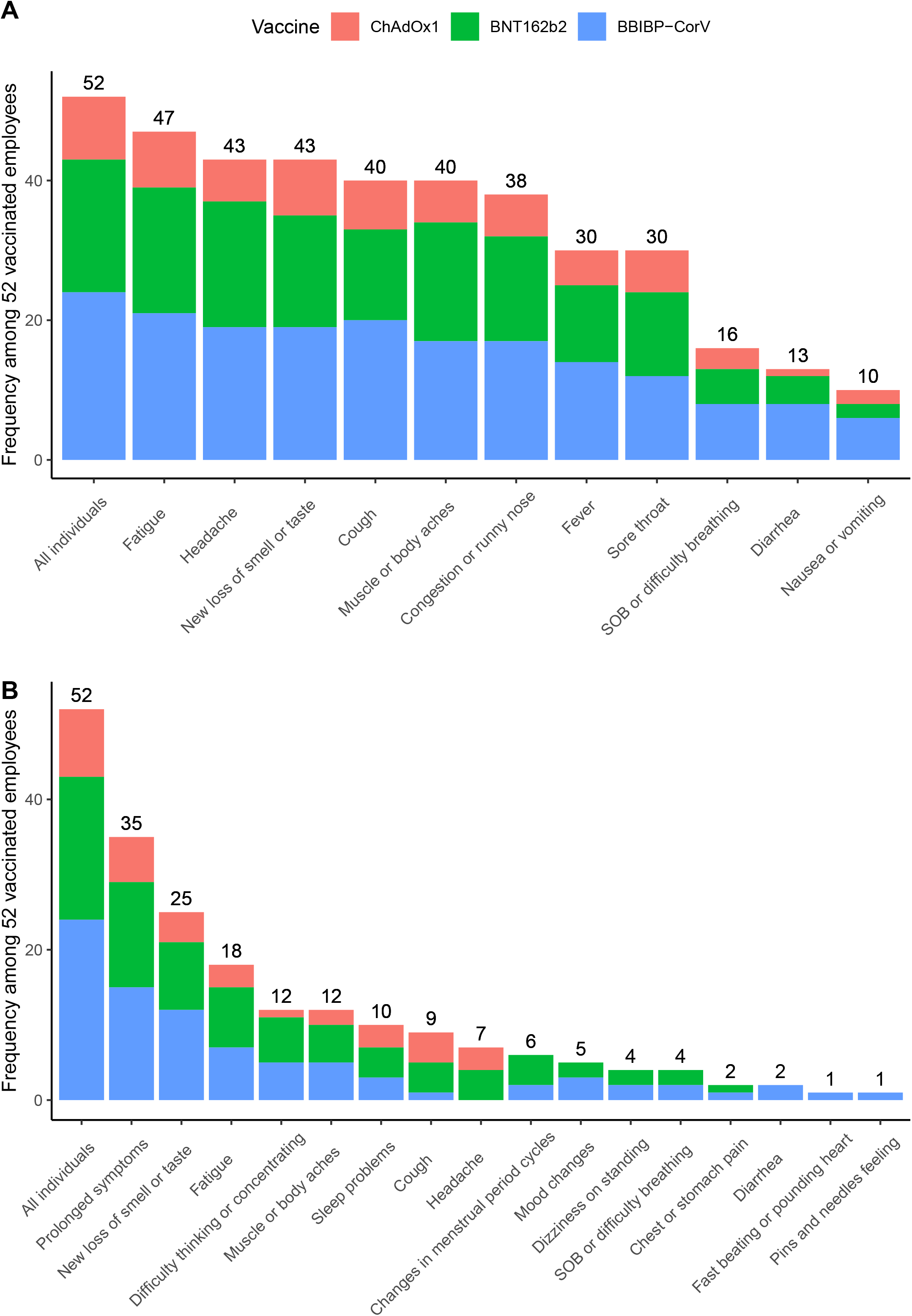
COVID-19 Symptoms Among 52 Vaccinated Health Care Workers, showing symptoms within the first month(A) and those lasting more than 28 days (B) after infection.

## Discussion

Our findings demonstrate the real-world differential efficacy of COVID-19 vaccines for protecting against PCR-confirmed SARS-CoV-2 infections. Despite the controversy in defining the efficacy endpoints of these emerging vaccines, we defined efficacy as the ability of the vaccines to prevent transmission as confirmed by PCR to avoid biases stemming from documentation of the symptoms and severity of disease.^6^ This strategy is best for studying relatively young populations, such as the HCWs included in our study, which may not experience the same level of disease severity as that in older populations.

The nature of the vaccination campaign in Jordan and its coincidence with an outbreak allowed for a great opportunity to compare the efficacy of inactivated whole-virus, mRNA and viral vector vaccines. Our institutional vaccination campaign yielded great success convincing HCWs to become vaccinated, with only 4.5% not receiving their first vaccine dose by the study cutoff date. This success resulted in part from annual campaigns requiring influenza vaccinations in prior years. Providing vaccinations to more than 1400 HCWs on campus also contributed to the success of the campaign. Our infection control staff and administrators also played an active role in disseminating knowledge throughout the institution.

We found that the BBIBP-CorV and ChAdOx1 vaccines were less effective in protecting against SARS-CoV-2 infections than was the BNT162b2 vaccine, which was predominantly used in Jordan. The BNT162b2 vaccine induces higher levels of antibodies and stronger cellular responses to SARS-CoV-2 than does BBIBP-CorV.^7^ A single dose of BNT162b2 also generates higher levels of SARS-CoV-2–specific antibodies than does ChAdIx1.^8^ This raises the important question of whether a booster vaccine with a different type of vaccination should be administered regularly to those who received inactivated whole-virus vaccines, particularly to those with no previous infections. We also observed great synergism between previous SARS-CoV-2 infection and vaccination for protecting against breakthrough infections, a finding previously reported for different types of COVID-19 vaccines.^9, 10^ How long this protection is afforded to these HCWs should be monitored to accurately determine the duration of efficacy from combined natural and vaccine-mediated immunity.

Real-world examples are helpful to improve our understanding of the COVID-19 vaccines, particularly when comparative trials are lacking.^11^ Despite the reported efficacy of inactivated whole-virus vaccines in preventing COVID-19,^12^ the countries relying on these vaccines have higher rates of reported infections than do countries that predominantly use other vaccine types.^13^HCWs are particularly vulnerable because of their high risk of repetitive exposures. The resurgence of infections among vaccinated HCWs is suggested to be dependent upon vaccine efficacy, time after vaccination and exposure to new strains.^14^

All contacted HCWs reported symptoms and two-thirds reported prolonged symptoms (> 28 days). In a multinational study reported by Sudre et al, 558 of 4182 patients with symptomatic COVID-19 had symptoms that lasted more than 28 days and 189 had symptoms lasting more than 56 days.^15^ We cannot rule out recall bias in our study because individuals are more likely to report symptoms in retrospective studies than in prospective studies. Our testing threshold may also have been biased towards symptomatic patients. Thus, some patients with very mild or asymptomatic disease may not have been included in our analysis.

The retrospective nature of our study introduced some obvious limitations. We cannot rule out some bias in selecting HCWs for specific types of vaccines. For example, our institutional campaign, which relied heavily on the BNT162b2 vaccine, was specifically directed towards our younger staff who were not prioritized to receive vaccines in the first few weeks of the national campaign.

In conclusion, high rates of COVID-19 vaccination can be achieved among HCWs, even in the presence of high rates of hesitancy. The BNT162b2 vaccine was superior to the other vaccines available in Jordan, suggesting its value as a booster vaccine for HCWs who previously received other vaccine types. Mixing and matching vaccines appears to be effective and safe,^16^ but much more research is needed.

## Data Availability

All data produced in the present study are available upon reasonable request to the authors

## Acknowledgement

Authors would like to thank Dr. Nisha Badders from St. Jude Children’s Research Hospital for helping in editing the final manuscript.

